# The effect of two community-based interventions on sanitation and hygiene outcomes in rural Malawi: a controlled before-and-after (CBA) trial

**DOI:** 10.1101/2025.09.30.25336959

**Authors:** Kondwani Chidziwisano, Clara MacLeod, Mindy Panulo, Blessings White, Joseph Wells, Ian Ross, Tracy Morse, Robert Dreibelbis

**Author notes:** Corresponding author: Kondwani Chidziwisano, Centre for Water, Sanitation, Health and Appropriate Technology Development (WASHTED), Malawi University of Business and Applied Sciences, Blantyre, Malawi. **Trial registration** The study was registered at clinicaltrials.gov (NCT05808218).

## Abstract

**Introduction:** Community Led Total Sanitation (CLTS) has been used to increase sanitation coverage and use. However, robust evaluations of CLTS with and without other community-based interventions remain limited. We evaluated the effectiveness of standard CLTS and CLTS combined with a community-based “Care Group” model on sanitation and hygiene outcome compared to a non-intervention control in Chiradzulu District, rural Malawi.

**Methods:** We conducted a three-arm controlled before-and-after trial. One arm received standard CLTS, one received CLTS plus Care Group (CLTS+CG), and one served as control. Baseline and endline surveys were administered to 1,400 randomly sampled households. Difference-in-difference analysis assessed changes in sanitation and handwashing outcomes between May-June 2023 (baseline) and April-May 2024 (endline).

**Results:** Access and use of sanitation facilities were generally high at baseline in all study arms. The primary outcome of the trial was access to a private sanitation facility of any quality. Both intervention arms were associated with improved odds of household having access to a private sanitation facility at endline compared to the control (CLTS: adjusted odds ratio [aOR] 3.11 (95% confidence interval [CI] 2.06–4.70); CLTS+CG: aOR 2.16 (CI 1.34–3.48)). However, there was no evidence that either intervention increased the odds of having a sanitation facility meeting quality standards for Sustainable Development Goals (SDG) 6. There was no observable difference between the two treatment arms on reported sanitation use. The odds of having a basic handwashing facility were higher in the CLTS+CG group versus control (aOR 2.62, CI 1.19–5.75) and CLTS+CG group versus standard CLTS group (aOR 2.89, CI 1.14–7.34); however, absolute increases in handwashing station coverage were limited.

**Conclusion:** This study suggests that CLTS is unlikely to result in improvements in sanitation of quality required to meet SDG targets and has minimal impact on household level investment in hygiene facilities. Standard CLTS should no longer be the *de facto* rural sanitation policy.

## Introduction

The Sustainable Development Goals (SDGs) have set the ambitious target of universal coverage of basic sanitation by 2030. At the domestic level, basic sanitation facilities are defined as sanitation facilities that are used by members of a single household and that are designed to maintain separation between excreta and human contact and are used by members of a single household (UNICEF & WHO, 2023). Globally, 1.5 billion people still lack access to basic sanitation – the quality level ascribed to in SDG 6 for water, sanitation, and hygiene (WASH) – with half of these people residing in sub-Saharan Africa (World Health Organization & UNICEF, 2025) Effective sanitation and hygiene interventions have the potential to reduce exposure to faecal pathogens and improve health (Pickering et al., 2019; Ziegelbauer et al., 2012) The global burden of disease could be reduced by preventing diarrhoeal diseases, soil-transmitted helminths and certain neglected tropical diseases linked to poor water sanitation and hygiene (WASH) (Wolf et al., 2023). In addition, access to improved sanitation can improve sanitation-related quality of life (Akter et al., 2025)and save travel time which has an economic value (Ross et al., 2025). The use of improved sanitation is also associated with lower antibiotic resistance as improved sanitation facilities prevents the load of antibiotic-resistant bacteria that could otherwise spread into the environment (Fuhrmeister et al., 2023).

Behaviour-centred interventions have been associated with improved uptake of sanitation and hygiene practices (Garn et al., 2017), but robust data focuses primarily on singular behavioural approaches. Community-Led Total Sanitation (CLTS) is one of the most widely used behaviour change interventions to improve household sanitation and hygiene behaviours in rural areas (Zuin et al., 2019). The approach works by catalysing collective community action to construct and use toilets as well as handwashing facilities through a series of community participation activities (Harter et al., 2020). Evaluations of changes in sanitation coverage following CLTS interventions are largely mixed, with noted reductions in open defaecation and increased latrine coverage in many cases (Garn et al., 2017; Venkataramanan et al., 2018). However, CLTS is less effective in areas where open defecation rates are already low and CLTS interventions rarely result in sanitation improvements that meet SDG targets (Crocker et al., 2016; Stuart et al., 2021). Recent reviews also report slippage back to open defecation following CLTS interventions (Kouassi et al., 2023).

Since its introduction in 2009, CLTS has evolved considerably by incorporating various subsidy programmes (Venkataramanan et al., 2018), market mechanisms, and other community-based behaviour change approaches. The incorporation of community-based structures into CLTS interventions can be effective for leveraging long-term behaviour change (Panulo et al., 2022; Ricca et al., 2014), but there is limited evidence on combined effect of CLTS with other community-based behaviour change strategies. The Care Group (CG) model is a community-based strategy for delivery of behaviour change interventions at community level (Weiss et al., 2015). Historically, focused on maternal and child health through a supportive network of peer-to-peer counselling (Davis et al., 2013; Perry et al., 2014), CG and their variants have been increasingly integrated into community-based WASH behaviour change interventions (Morse et al., 2020; Panulo et al., 2022). CGs can extend the reach of behaviour change interventions by providing additional points of contact with target households and groups. Although the CG model has been tested with successful results in community-based interventions for maternal health and infant and young child nutrition and feeding (Chidziwisano et al., 2020; Morse et al., 2020), there is little evidence on its effectiveness for promoting broader household sanitation and hygiene behaviours. It is also unknown to what extent combining the CG model with existing community-led WASH interventions, such as CLTS, can have additive or synergistic impact on sustained sanitation coverage and use.

This study aims to assess the effectiveness of a CLTS intervention with and without a CG model on sanitation outcomes in rural Chiradzulu District of Malawi. The study objectives were to assess: 1) whether the two interventions are individually more effective than no intervention at all, and 2) how the two interventions compare to each other.

## Methods

Full details of the study design, setting and intervention, for the larger trial are reported elsewhere (Chidziwisano et al., 2025).

### Study design

This study was a controlled before-and-after (CBA) trial of the “Water, Sanitation and Hygiene for Everyone” (W4E) programme implemented by World Vision and Water for People. The programme targeted universal access to WASH in Chiradzulu District, Malawi by 2024. The intervention was delivered at Traditional Authority (TA) level and included three arms: i) CLTS only, ii) CLTS+CG, and iii) control. Both intervention arms received the assigned intervention activities for 11 months, while the control arm received no intervention.

### Study setting and population

The study took place in Chiradzulu District, located in the southern region of Malawi (Figure 1a). The District is sub-divided into 10 administrative regions, also known as Traditional Authorities (TA) (Malawi Government, 2023). The trial was conducted in three TAs: Chitera, Ntchema, and Nkalo (Figure 1b), which are predominantly rural. TAs Chitera and Ntchema were randomly allocated to the CLTS+CG and CLTS only arms, respectively, while TA Nkalo was randomly allocated as the control group. Households in each treatment arm were randomly selected for the trial.

**Figure 1.**
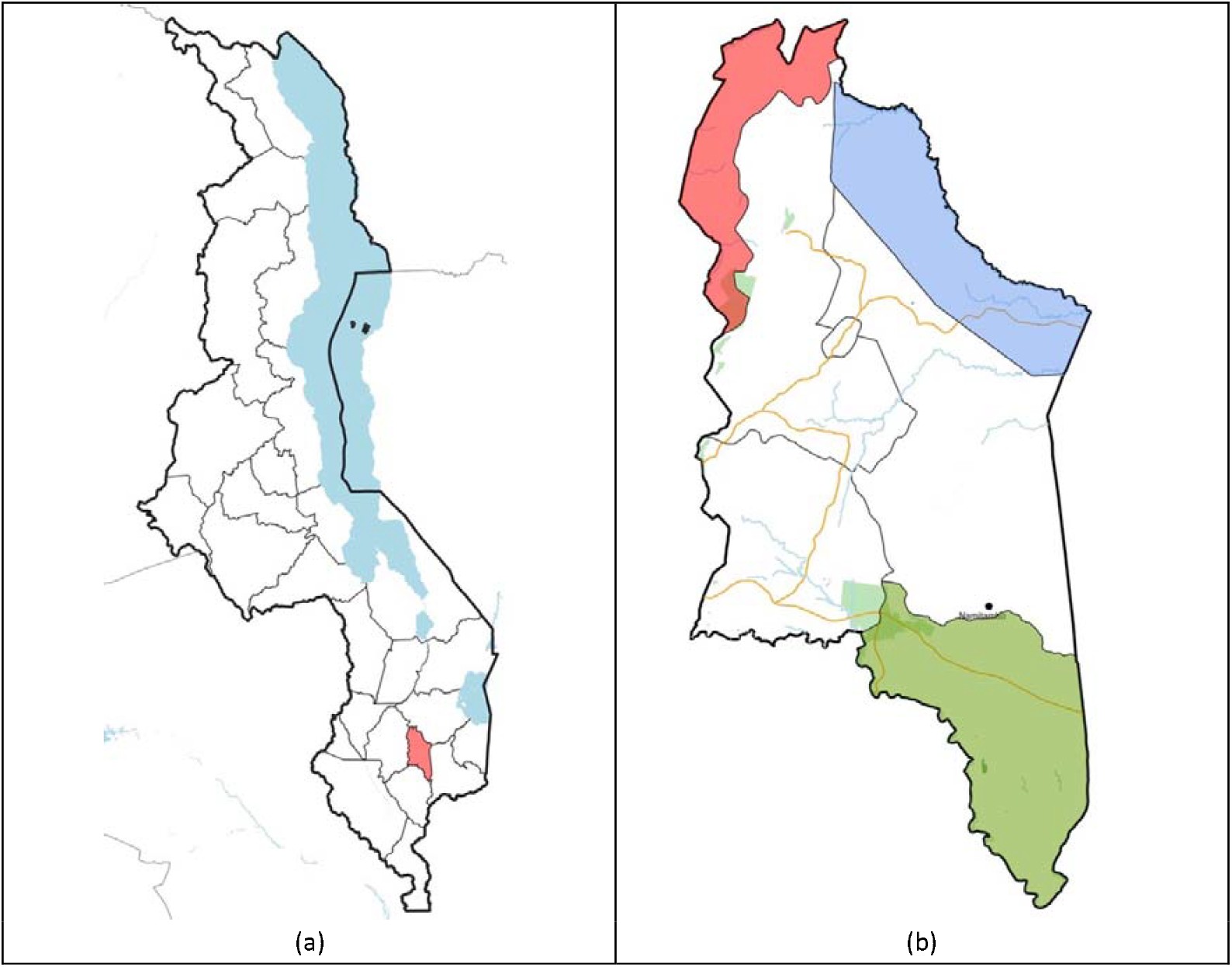
(a) Map of Malawi with Chiradzulu District highlighted in red; (b) Map of Chiradzulu District, Malawi, with TA Chitera (CLTS+CG) highlighted in red, TA Ntchema (CLTS) highlighted in blue, and TA Nkalo (control) highlighted in green. The base layer was obtained from Database of Global Administrative Areas (GADM) (available from: https://gadm.org/download_country.html). The terms of use can be found here: https://gadm.org/license.html. Road and waterbody features were obtained from OpenStreetMap under the Open Database License (available from: https://www.openstreetmap.org).

**Figure 2.**
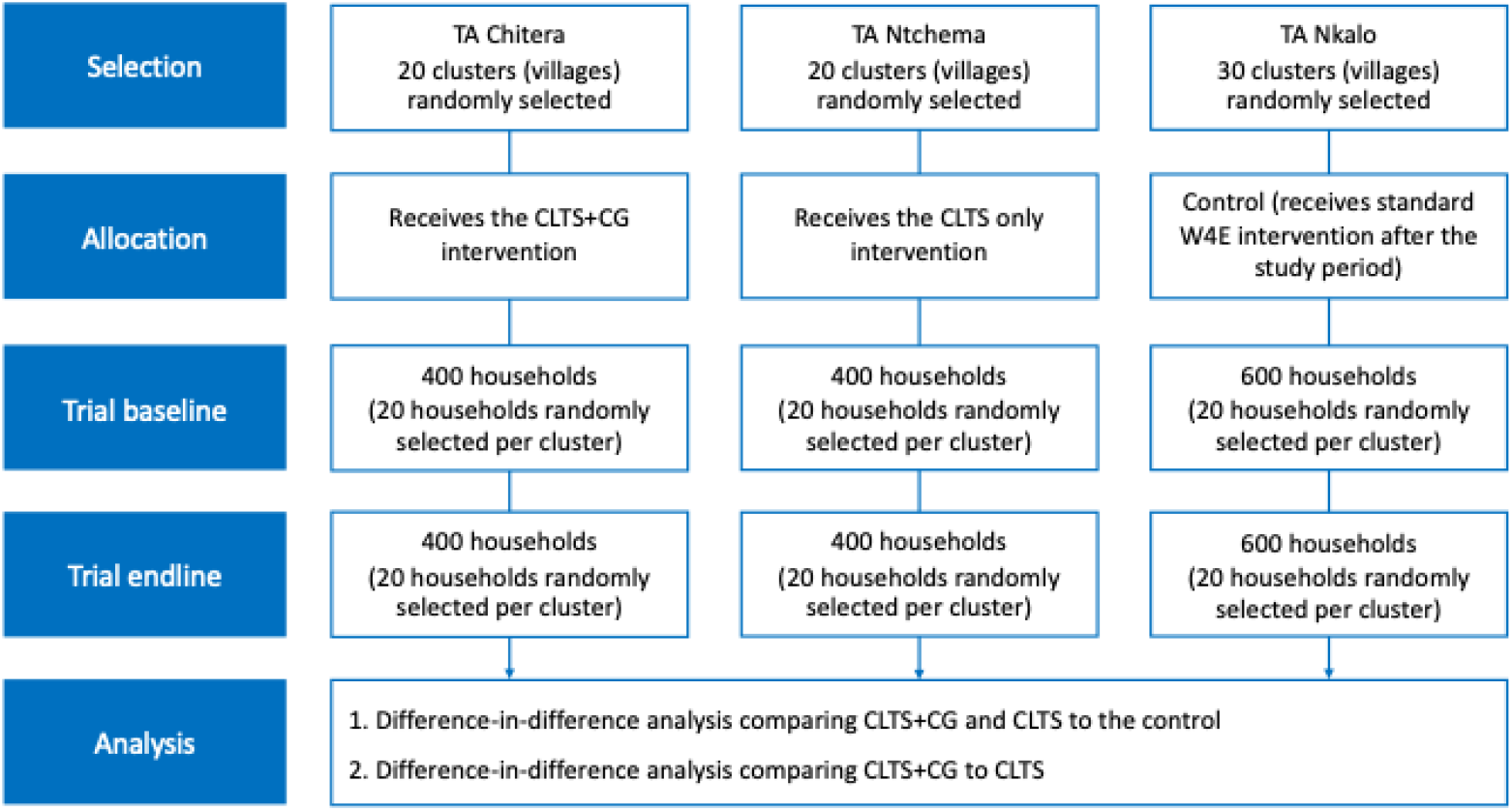
Participant flow diagram.

### Description of the intervention

The full intervention is described elsewhere (Chidziwisano et al., 2025), but the main sanitation and hygiene intervention components are described here.

#### CLTS

The CLTS intervention arm (TA Ntchema) received a standard CLTS intervention, delivered according to the Malawi National Sanitation and Hygiene Strategy (Government of the Rublic of Malawi, 2018) and CLTS Handbook (Kar & Chambers, 2008). The CLTS activities were delivered in three phases: pre-triggering, triggering, and household follow ups. The follow up visits also included messages to promote sanitation coverage and proper use of sanitation facilities. In addition, the District Health Officers implemented hygiene promotion campaigns promoting handwashing with soap and building an improved handwashing station in the home.

#### CLTS+CG

The CLTS+CG arm (TA Chitera) received the same CLTS intervention as the CLTS arm. The CLTS+CG arm received additional village-level sanitation promotion through locally established care groups. CGs were designed to extend the reach of CLTS behaviour change messaging, providing additional points of contact with intervention households. Each care group consisted of one promoter and approximately 10 community health volunteers known as cluster leaders responsible for regularly visiting their beneficiary households and providing advice on sanitation. CG promoters and cluster leaders supported intervention delivery by holding participatory care group meetings with households. CGs also conducted CLTS follow-up visits to households in their communities, tracking progress against sanitation coverage and proper use of sanitation facilities.

#### The control

The control arm (TA Nkalo) did not receive any intervention but received the CLTS intervention after data collection was completed in the two intervention TAs.

### Selection of clusters and households

Clusters were villages within the targeted Traditional Authorities (TAs). Household and cluster selection methods are detailed elsewhere (Chidziwisano et al., 2025). In each selected village, systematic sampling methods were used to select 20 households for enrolment.

### Data collection

Baseline data was collected in April and May 2023 prior to W4E implementation in the CLTS and CLTS+CG arms. Endline data collection was conducted in March 2024 before CLTS implementation in the control arm. A structured questionnaire with close-ended questions and pre-coded responses was used to collect data on household characteristics and sanitation and hygiene outcomes. The household survey was conducted in Chichewa, the local language, by 10 trained enumerators. Data was collected using mobile devices and uploaded directly online to KOBO daily. Only the PI, co-investigators, and study personnel had authorised access to the data.

### Study Outcomes

The primary outcome of this study was sanitation coverage, defined as households having a sanitation facility of any quality that was not shared with other households (Table 1). Secondary outcomes included household sanitation coverage that met the JMP criteria basic sanitation facility or better (UNICEF & WHO, 2023), individual sanitation use (self-reported), and presence of a handwashing facilities in the home (Table 1). Handwashing facilities – a dedicated location and/or mobile device used for washing hands – were classified as basic (having both soap and water available at time of data collection), limited (having either soap or water but not both) or no facility (UNICEF & WHO, 2023). Outcomes were measured via an interviewer-administered questionnaire at baseline and 11 months after intervention delivery. Sanitation use was measured for all individuals present at the time of data collection. All other outcomes were measured at the household level based on a primary respondent.

**Table 1.** Description of the study outcomes.

| Outcomes | Description |
| --- | --- |
| Sanitation coverage | A household having a private sanitation facility of any quality |
| Basic sanitation coverage | A household-level variable based on presence of a private improved sanitation facility as observed by a fieldworker, (designed to hygienically separate excreta from human contact) as defined by the JMP (UNICEF & WHO, 2023) |
| Sanitation use | An individual-level binary variable based on whether the respondent reported using a sanitation facility at the time of the last defaecation event. |
| Basic handwashing facility | A household-level binary variable based on the presence of a handwashing facility with both soap and water as defined by the JMP (UNICEF & WHO, 2023), fieldworker-observed in most cases. |

### Data analysis

A household-level binary variable based on the presence of a handwashing facility with both soap and water as defined by the JMP (UNICEF & WHO, 2023), fieldworker-observed in most cases. Data was cleaned using R statistical software (R Studio Team, 2021) and analysed using Stata v18 (Stata Corp, College Station, TX). The analysis used an intention to treat approach where data was analysed according to the treatment arm (Hollis & Campbell, 1999). A difference-in-difference (DID) analysis was used to estimate the intervention effect adjusted for baseline differences in basic sanitation coverage and sanitation use. The DID models were estimated using multilevel mixed-effects logistic regression with robust standard errors to account for clustering (melogit). Regression coefficients were converted to odds ratios. For each sanitation outcome, we reported the odds ratio, 95% confidence interval, and p-value for both treatment arms compared to the control, as well as for the CLTS+CG group compared to the CLTS group.

The first set of models assessed differences in outcomes with adjustment only for clustering and for community size (above or below the TA-specific median), a design variable. A second set of models, which we refer to as our primary analysis, adjusted for pre-specified covariates including number of household members, at least one household member with a disability, wealth quintile, water source on plot, and village size. The wealth index was constructed by combining information on household asset ownership among households at baseline using principal components analysis(Vyas & Kumaranayake, 2006) and converting the asset index into quintiles. Asset weights from baseline were applied to household asset information at endline and converted to quintiles.

## Ethics

This study received prior approval from the National Commission for Science and Technology (P01/23/718) and by the London School of Hygiene and Tropical Medicine Research Ethics Committee (Ref: 28249). Further, consent to conduct the study was obtained from Chiradzulu District Council and local leaders. All study participants provided written informed consent prior to data collection. In case of an illiterate participant, any adult household member (18 years and above) was asked to sign on behalf of the participant as a witness.

## Results

1,400 households were included at both baseline and endline, with information on sanitation use available for the majority of the individuals (Tables 2 and 3). Overall, no major imbalances were observed between the arms at baseline (Chidziwisano et al., 2025) and endline (Table S1).

**Table 2.**
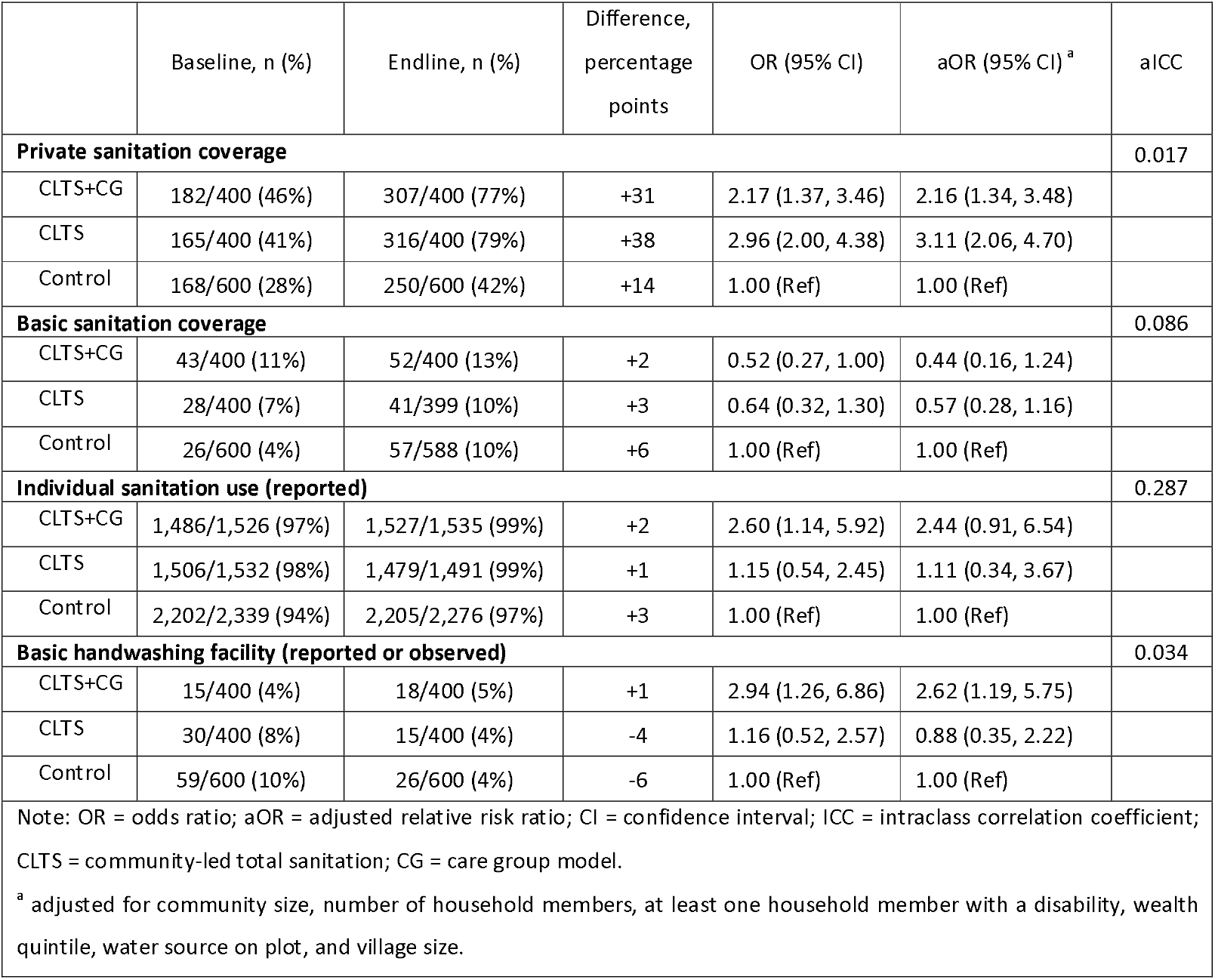
CLTS and CLTS+CG vs control for the trial outcomes.

**Table 3.** CLTS+CG vs. CLTS for the trial outcomes.

|  | Baseline, n (%) | Endline, n (%) | Difference, % | OR (95% CI) | aOR (95% CI) <sup>a</sup> |
| --- | --- | --- | --- | --- | --- |
| **Sanitation coverage** |  |  |  |  |  |
| CLTS+CG | 182/400 (46%) | 307/400 (77%) | +31 | 0.73 (0.43, 1.27) | 0.70 (0.39, 1.24) |
| CLTS | 165/400 (41%) | 316/400 (79%) | +38 | 1.00 (Ref) | 1.00 (Ref) |
| **Basic sanitation coverage** |  |  |  |  |  |
| CLTS+CG | 43/400 (11%) | 52/400 (13%) | +2 | 0.80 (0.41, 1.58) | 0.78 (0.29, 2.05) |
| CLTS | 28/400 (7%) | 41/399 (10%) | +3 | 1.00 (Ref) | 1.00 (Ref) |
| **Individual sanitation use (reported)** |  |  |  |  |  |
| CLTS+CG | 1,486/1,526 (97%) | 1,527/1,535 (99%) | +2 | 2.26 (0.80, 6.38) | 2.11 (0.56, 8.01) |
| CLTS | 1,506/1,532 (98%) | 1,479/1,491 (99%) | +1 | 1.00 (Ref) | 1.00 (Ref) |
| **Basic handwashing facility (reported or observed)** |  |  |  |  |  |
| CLTS+CG | 15/400 (4%) | 18/400 (5%) | +1 | 2.52 (0.98, 6.49) | 2.89 (1.14, 7.34) |
| CLTS | 30/400 (8%) | 15/400 (4%) | -4 | 1.00 (Ref) | 1.00 (Ref) |
| Note: OR = odds ratio; aOR = adjusted relative risk ratio; CI = confidence interval; CLTS = community-led total sanitation; CG = care group model. |  |  |  |  |  |
| <sup>a</sup> adjusted for community size, number of household members, at least one household member with a disability, wealth quintile, water source on plot, and village size. |  |  |  |  |  |

### Sanitation coverage

Changes in sanitation coverage between baseline and endline for each of the three study areas are presented in Figure 3. Across all groups, almost all households had access to a sanitation facility of some type at baseline, and shared sanitation facilities – both limited facilities and shared unimproved facilities – were common in all three treatment arms. In all three groups we see a notable decrease in the use of shared sanitation facilities and an increase in private unimproved facilities between baseline and endline data collection.

**Figure 3.**
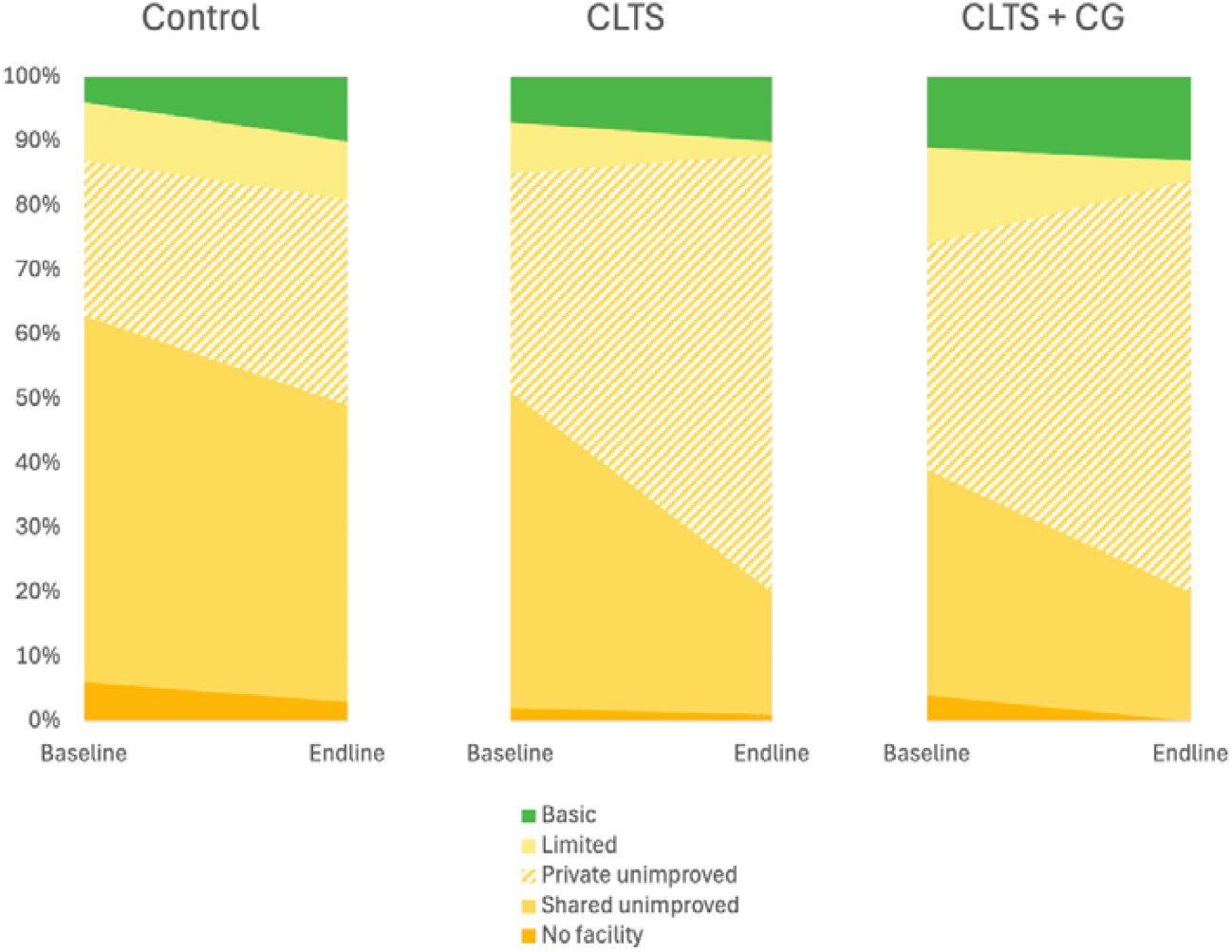
Sanitation coverage according to the JMP sanitation ladder (UNICEF & WHO, 2023). The levels of service were defined as i) basic (use of an improved facility that is not shared with other households), ii) limited (use of an improved facility that are shared with other households), iii) unimproved (use of a pit latrine without a slab or platform, hanging toilet or bucket latrine), and iv) no sanitation facility (also defined as open defecation, or the disposal of faeces in the environment such as in fields or forests). We disaggregated unimproved facilities as either shared (between at least two households) or private.

In our difference-in-difference analysis, households in the CLTS arm had approximately three times greater odds of having a private sanitation facility of any type compared to the control arm (adjusted Odds Ratio [aOR] = 3.11, 95% confidence interval [CI]= 2.06, 4.70) and two times higher odds in the CLTS+CG group (aOR = 2.16, CI = 1.34, 3.48). In contrast, we found no evidence that either intervention was associated with a change in the odds that a household would use a basic or better sanitation facility compared to the control group (CLTS aOR = 0.57, CI 0.28, 1.16; CLTS+CG aOR = 0.44, CI = 0.16, 1.24).

Comparing the two treatment arms, we found no evidence of a difference in the odds of having private a sanitation facility of any quality between the two interventions (aOR 0.70, CI = 0.39, 1.24). We found no evidence of a difference in the odds of having a basic sanitation facility in the CLTS+CG group compared to the CLTS group (aOR = 0.78, CI = 0.29, 2.05).

### Sanitation use

Reported sanitation use at baseline was high in all three study groups (Table 2) and remained high at endline (Table 2 and 3). At the individual-level, we found no evidence of differences in the odds of reported sanitation use (in the last time one defaecated) among individuals in the CLTS group (aOR = 1.04, CI = 0.32, 3.40) or the CLTS+CG group compared to the control group (aOR = 2.53, CI = 0.97, 6.56). We found no evidence of a difference in the odds of individual sanitation use in the CLTS+CG group compared to the CLTS group (aOR = 2.37, CI = 0.62, 9.03) (Table 3).

### Presence of a basic handwashing facility

Across study arms, access to a basic handwashing facility did not exceed 10% in any study arm (Figure 4) and coverage declined in all study groups between baseline and endline. Between 45% and 50% had limited handwashing facilities, defined as dedicated location or device but lacking either water or soap at the time of data collection, at both baseline and endline.

**Figure 4.**
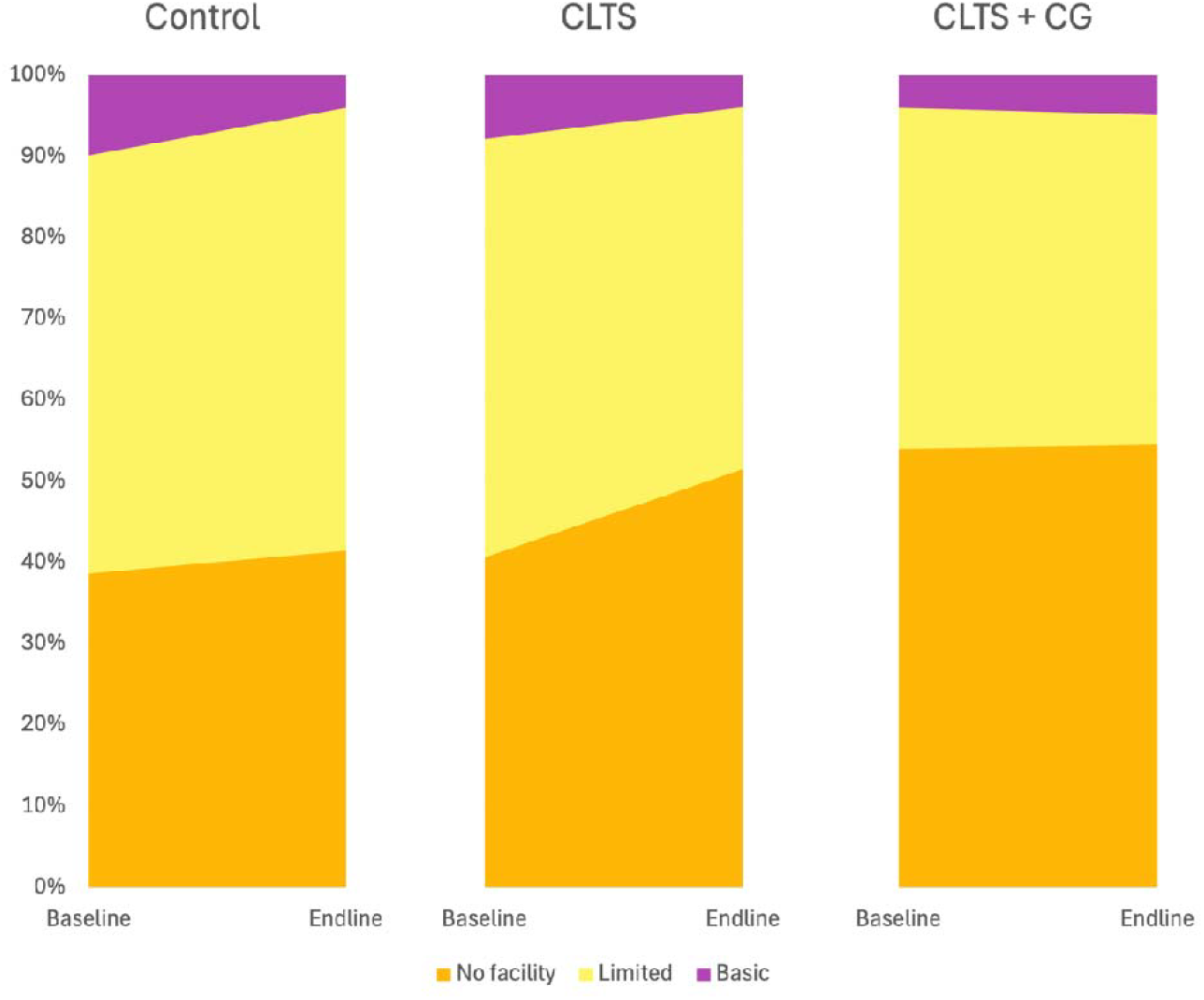
Observed or reported handwashing facility coverage at baseline and endline in each study arm. Basic handwashing facility is defined as the availability of a handwashing facility with soap and water at home. Limited as the availability of a handwashing facility lacking soap and/or water at home. No facility is defined as no handwashing facility on premises.

We found no evidence of differences in the odds of a household having an observed or reported basic handwashing facility among households in the CLTS group and the control group (aOR = 0.88, CI = 0.35, 2.22). However, households in the CLTS+CG group had 2.6 great odds of having a basic handwashing facility compared to the control arm (aOR = 2.62, CI = 1.19, 5.75). We also observed increased odds of having a basic handwashing facility in the CLTS+CG group compared to the CLTS group (aOR 2.89, CI = 1.14, 7.34, p=0.026) (Table 3).

## Discussion

This study investigated the effect of a CLTS intervention with and without CG model on sanitation and hygiene outcomes in rural Chiradzulu District, Malawi. Baseline sanitation coverage of any quality was high in our study area, with large portions of the population using shared facilities. Our study found significant increases in access to a private sanitation facility of any quality in both intervention arms when compared to a control arm, but no observable difference in access to basic sanitation facilities. Interventions appeared successful in helping people make the transition from shared unimproved to private unimproved facilities, but limited incremental improvement in sanitation quality (Figure 3). Use of a sanitation facility at the last defaecation event was over 95% in our study population at baseline and increased in all study arms. Neither intervention was associated with large changes in access to an improved handwashing facility.

We note limited additional benefits associated with CLTS with embedded Care Group support compared to CLTS alone, despite the fact that Care Group models have proven effective in improving health behaviours in other community-based health programmes in similar context (Morse et al., 2020). An embedded process evaluation of the two intervention arms in this trial found that Care Groups were engaged primarily as an extension of monitoring CLTS rather than enhancing its implementation, so were underutilised (Panulo, et al., 2025). Their role in intervention delivery and follow-up was focused on documenting household sanitation construction, a task already covered by natural leaders and community health workers (i.e. Health Surveillance Assistants) in the CLTS only arm. In contrast, traditional Care Group interventions provide direct one-to-one support to households and new mothers around adoption of key child health behaviours (Panulo et al., 2022). As many environmental health interventions are increasing their reliance on Care Groups or analogous community-based organisations, our study suggests that engagement requires intentional planning and coordination to ensure that the Care Groups provide an additive, complementary intervention rather than just task shifting.

Studies have shown that standard CLTS interventions mainly focus on sanitation promotion with minimal efforts towards hygiene activities (Biran et al., 2020; Whaley & Webster, 2011). The lack of improvements in both handwashing facilities and observed handwashing behaviour (Macleod, 2025) suggest the hygiene promotion component of both standard and modified CLTS programming requires significant strengthening. The Care Group model, which provides direct support to households in adopting healthy behaviours (Morse et al., 2020; Panulo et al., 2022) provides a potential natural extension to the reach of large-scale sanitation interventions and warrants further exploration in these settings. In our study however, Care Groups did not appear to have an incremental benefit for handwashing assets or behaviour.

The high rates of sanitation access at baseline and low rates of reported defaecation suggest that CLTS may not be fit for purpose in Malawi. CLTS, as originally conceived, aimed to end open defaecation irrespective of sanitation facility quality (Kar & Chambers, 2008). Since its emergence in the early 1990s, CLTS has become national policy in Malawi and many other countries. Like other studies (Harter et al., 2020; Kouassi et al., 2023; Laura et al., 2015), our results show CLTS and CLTS+CG interventions were associated with large observed increased in unimproved facilities but little to no change in higher quality sanitation coverage. Similar to our study, CLTS has been shown to be ineffective where open defaecation rates are already low (Crocker et al., 2016; Stuart et al., 2021). In its current form, CLTS will likely result in limited progress against Malawi’s 2030 Malawi Implementation Plan 1 (MIP 1) (Government of Malawi, 2021) and the Sustainable Development Goals (SDGs) targets (Hinton et al., 2023). In settings with low levels of open defecation, interventions that more effectively target improvements in sanitation quality rather than just coverage are needed, along with rigorous evaluation of their effectiveness.’’

CLTS-based sanitation programming is unlikely to achieve the full potential of sanitation interventions to maintain healthy environments. Unimproved sanitation facilities, contribute to the transmission of enteric pathogens, such as Escherichia coli (*E. coli*) in the broader environment (Gizaw et al., 2022). Unsafe containment of faecal sludge that leaks and/or overflows into nearby drains or other surface water bodies also contributes to environmental dispersion of antimicrobial resistant pathogens, as evidenced across Malawi and Uganda (Musicha et al. 2025, Cocker, et al. 2023; Mwapasa et al., 2024). Preventing the transmission of enteric pathogens through improved environmental health infrastructure is therefore essential for protecting public health (Musicha, et al., 2024; Mwapasa et al., 2024) (Fuhrmeister et al., 2023). These risks are compounded by the poor behavioural and physical sustainability of CLTS interventions. In Malawi, Hinton and colleagues have found high rates of attrition following CLTS interventions (Hinton et al., 2024). Several studies have shown a high risk of collapse of unimproved facilities during the rainy season and extreme weather events (Cole, 2015; Hinton et al., 2024; MacLeod et al., 2024; Nijhawan et al., 2025). Climate change will further compound the risks posed to sanitation infrastructure, and intervention models that go beyond unimproved facilities while explicitly focusing on climate-resilient sanitation services are needed.

Despite its limitation, there are few alternatives to CLTS programming that can be readily integrated into policy and programming. National-level programmes in India (Curtis, 2019) and Tanzania (Aunger et al., 2024) have been associated with both increased demand for and measured improvements in improved sanitation, but these interventions required significant national commitment, stakeholder coordination, and financing (Mehta, 2018). Implementation at the national level makes evaluation and understanding the mechanism of impact difficult. Market-based approaches have been increasingly proposed as possible intervention models for improving sanitation quality. The equity implications of these approaches require further research and there have been limited robust evaluations of their effectiveness (O’Keefe et al., 2015). More experimentation and coordinated efforts are needed to identify locally appropriate and effective models for sanitation improvements.

This study has limitations. All data about sanitation coverage and use were self-reported. Confirmation of self-reported data was conducted through fieldworker observations of sanitation and handwashing facilities reported elsewhere (Macleod et al, 2025). The follow-up period was limited to only 11 months; the study results do not reflect the long-term sustainability of the CLTS and CLTS+CG interventions. Given district-level implementation, we were unable to randomize treatment arms at the household or village-level. We have attempted to control for possible confounders but cannot rule out the possibility of residual confounding in our results. The magnitude of changes in private sanitation coverage in our intervention communities and the lack of observed changes in handwashing facility coverage suggests that the role of confounding in our study may be minimal. Blinding of the intervention to participating households was not possible. However, the data collectors were blinded to the treatment allocation.

## Conclusion

It is time for national governments to reconsider current sanitation policy and end the hegemony of CLTS as the *de facto* approach for improving sanitation. More context specific interventions reflective of local sanitation coverage and use are needed to make true progress on national and global sanitation targets. Achieving the environmental health benefits of sanitation interventions requires intervention models that go beyond unimproved sanitation facilities and focus on basic or safely managed systems. The modified version of Care Group model used in the WASH for Everyone Project was insufficient to bring additional benefits in sanitation and hygiene outcomes, and more innovative and deliberate integration of the Care Group approach into other community-based programming is needed.

## Data Availability

All data produced in the present study are available upon reasonable request to the authors

## Acknowledgements

We would like to extend our gratitude to all the participating households who allowed us into their homes and made this study possible. Thank you also to the enumerators for their hard work collecting the data.

## Funding

This work was supported by World Vision US (grant number WVSOW34730 to LSHTM).

## Conflict of interest

No conflict of interest

## Supplementary materials

**Table S1.**
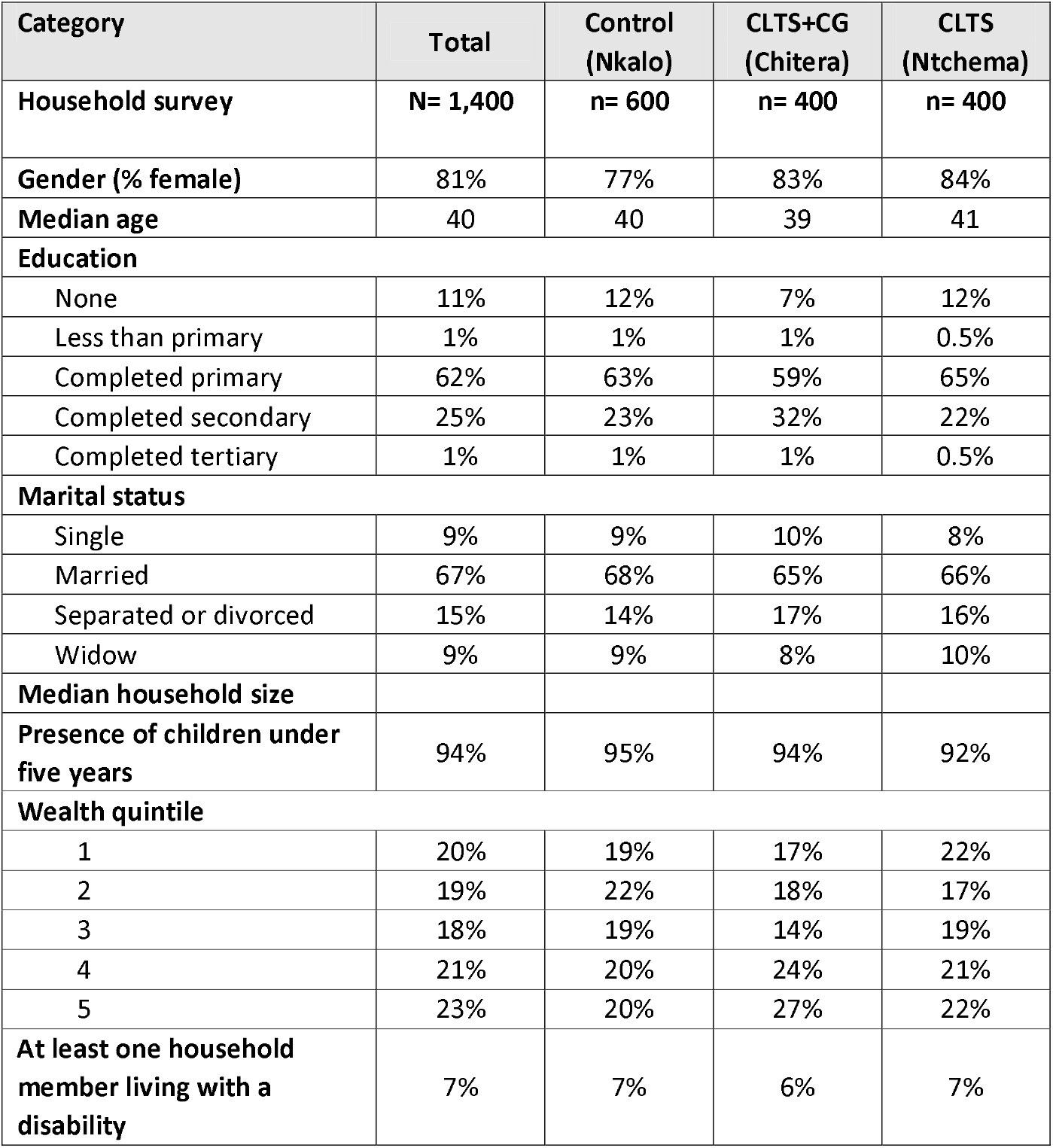
Individual and household characteristics of participants at endline.

## Notes

### Competing Interest Statement

The authors have declared no competing interest.

### Clinical Trial

NCT05808218

### Funding Statement

This study was funded by World Vision US (grant number WVSOW34730 to LSHTM)

### Author Declarations

Ethics committee/IRB of NATIONAL COMMITTEE ON RESEARCH IN THE SOCIAL SCIENCES AND HUMANITIES gave ethical approval for this work.

